# Plasma P-tau 181, P-tau 217 and Other Blood-Based Alzheimer’s Disease Biomarkers in a Multi-Ethnic, Community Study

**DOI:** 10.1101/2020.09.10.20192146

**Authors:** Adam M. Brickman, Jennifer J. Manly, Lawrence S. Honig, Danurys Sanchez, Dolly Reyes-Dumeyer, Rafael A. Lantigua, Patrick J. Lao, Yaakov Stern, Jean Paul Vonsattel, Andrew F. Teich, David Charles Airey, Nicholas Kyle Proctor, Jeffrey L. Dage, Richard Mayeux

**Author notes:** Correspondence: Richard Mayeux, MD Neurological Institute 710 West 168^th^ Street New York, NY 10032 (T).

## Abstract

**Introduction:** Blood-based Alzheimer’s disease (AD) biomarkers provide opportunities for community studies and across ethnic groups. We investigated blood biomarker concentrations in the Washington Heights, Inwood, Columbia Aging Project (WHICAP), a multi-ethnic community study of aging and dementia.

**Methods:** We measured plasma Aβ40, Aβ42,T-tau, P-tau181 and P-tau217, and neurofilament light chain (NfL) in 113 autopsied participants, (29% with high AD neuropathological changes) and in 300 clinically evaluated individuals (42% with clinical AD). Receiver operating characteristics were used to evaluate each biomarker. We also investigated biomarkers as predictors of incident clinical AD.

**Results:** P-tau181, P-tau217 and NfL concentrations were elevated in pathologically and clinically diagnosed AD. Decreased Aβ42/Aβ40 ratio and increased P-tau217 and P-tau181 were associated with subsequent AD diagnosis.

**Discussion:** Blood-based AD biomarker concentrations are associated with pathological and clinical diagnoses and can predict future development of clinical AD, providing evidence that they can be incorporated into multi-ethnic, community-based studies.

## 1. Introduction

Alzheimer’s disease (AD), the leading cause of dementia, is characterized by progressive impairment of memory and other cognitive functions resulting from neuronal loss, gliosis, extracellular amyloid deposits, and neurofibrillary tangles^1^. In 1984^2^, the NINCDS-ADRDA diagnostic criteria for AD, categorized three diagnostic levels: definite AD (neuropathological diagnosis), probable AD (clinical diagnosis with minimal confounding factors), and possible AD (clinical diagnosis with comorbidities). The sensitivity and specificity of the clinical criteria for probable AD compared with the postmortem diagnosis was of 81% and 70%, respectively^3^. In 2011^3^, the criteria were revised to recognize the pathological process that begins prior to the onset of clinical symptoms. The use of magnetic resonance imaging (MRI), positron emission tomography (PET) imaging, and cerebrospinal fluid (CSF) assays were included more systematically into diagnosis. Additional sets of criteria, one for preclinical AD^4^ and the other for mild cognitive impairment (MCI) due to AD^4^, were introduced that explicitly incorporated evidence of positive biomarkers (MRI, PET and CSF) into diagnoses. By 2018, the diagnostic scheme^5^ based on inclusion of biomarker evidence of amyloid (’A’), tau (’T’), and neurodegeneration (’N’) ^6^, and was recommended.

In the clinical setting, incorporating biomarkers into the diagnosis of AD is somewhat easier compared with incorporating them into observational studies that include thousands of individuals. The widespread use of PET and CSF biomarkers is difficult because of limitations in access to radiopharmaceuticals and performing lumbar punctures, respectively. Recent developments of AD blood-based biomarkers ^7-10^ may overcome these issues and provide an opportunity to derive diagnostic information with improved accuracy in large-scale observational research.

Here we used stored plasma to measure blood-based biomarkers in a multi-ethnic community-based study where diagnosis of AD was defined either clinically or neuropathologically. Both the clinical and autopsy group consisted of individuals from the Washington Heights, Inwood and Columbia Aging Project (WHICAP) cohort. Our focus was on the current state-of-the-art AD-related plasma biomarkers, including Aβ40 and Aβ42 as markers of amyloid pathology, total tau and Neurofilament Light (NfL) chain as markers of neurodegeneration, and Phospho-tau (P-tau) 181 and 217 as markers of tau pathology. We compared plasma biomarker concentrations between clinically- and pathologically-defined diagnostic groups and examined differences by race/ethnicity groups. A subset had undergone florbetaben PET to assess cortical Aβ plaque burden. We examined the relationship between blood-based biomarker concentrations and cerebral amyloidosis.

## 2. Methods

### 2.1 Participants

We selected all individuals (n=113) from WHICAP who had brain autopsy with pathological examination and had stored plasma. We also selected 300 individuals from the clinical cohort for analysis; the goal was to include equal numbers of participants from each of the three major race/ethnicity groups represented in WHICAP with relatively equal numbers of individual characterized at having clinical AD at their last available diagnostic visit. Race/ethnicity was self-reported based on the 2000 US census classification^11^: Non-Hispanic White (White), Hispanic, and non-Hispanic Black/African American. Within each group, participants were considered for inclusion if they had stored plasma for biomarker assays and had been assessed clinically on more than one occasion. All participants received a neuropsychological test battery, a structured medical and neurological examination, and blood sampling at study entry and at 18 to 24 month intervals. An independent consensus committee derived diagnoses of clinical AD^3^, control or other forms of dementia. The diagnosis of clinical AD included individuals with frank dementia and those with a Clinical Dementia Rating (CDR^12^) score of 0.5 deemed by the consensus committee as having a clinical syndrome consistent with very early or mild AD. For primary analyses in the clinical sample, we compared individuals with and without clinical AD at the time of the blood draw used to derive biomarker concentrations. For those without AD, we investigated the last available follow-up diagnosis to compare individuals who subsequently developed clinical AD with those who remained unimpaired. A subset of the clinical cohort (n=40) had received Florbetaben PET scanning. Informed consent was obtained from all participants.

### 2.2 Plasma Aβ42 and Aβ40, total tau and NfL

Centrifuged plasma aliquoted in polypropylene tubes and stored at -80°C were used to measure Aβ42, Aβ40, and total-tau using SIMOA technology (Quanterix, Lexington, MA, USA). The multiplex Neuro 3-plex A kit (#101995), and NfL kit (#103400) were used on 96-well plates. Rapid-thawed plasma (25 μL diluted 4-fold in buffer) was added to kit beads (100 μL) by pipette in each well, the plate was incubated for 15 min at 30°C shaking at 1000 rpm, magnetic-washed 3x for 5 min total, subjected to addition of SBG reagent (100 μL), followed by another incubation for 10 min at 30°C at 1000 rpm, washed again x 5 for 7 min total and read on the SIMOA SR-X machine (Quanterix). Each plate assays – in duplicate – 34 samples, 8 calibrators and 2 controls. We considered the ratio of Aβ42 to Aβ40 (Aβ42/Aβ40) as the primary amyloid biomarker.

### 2.3 Plasma P-tau181 and P-tau217

The P-tau assays were optimized to measure disease-related differences through the selection of monoclonal antibodies. Selection of the monoclonal antibody pair provided a unique combination of sensitivity and selectivity for the tau forms in plasma that differ between AD and HC participants. The P-tau181 assay has been modified from that published previously^8, 9^ in order to improve the assay and more directly compare between phosphorylation sites (see supplemental methods for additional details). The assays were performed on a streptavidin small spot plate using the Meso Scale Discovery (MSD) platform. For the P-tau181 assay, Biotinylated-AT270 was used as a capture antibody (anti-P-tau181 antibody) and SULF0-TAG-Ru-4G10-E2 (anti-tau monoclonal antibody) for the detector. The assay was calibrated using a synthetic P-tau181 peptide. For the P-tau217 assay, Biotinylated-IBA493 was used as a capture antibody (anti-P-tau217 antibody) and SULFO-TAG-Ru-4G10-E2 (anti-tau monoclonal antibody) for the detector. The assay was calibrated using a synthetic P-tau217 peptide.

### 2.4 Autopsy

Cases were classified diagnostically according to the National Institute on Aging-Alzheimer’s Association (NIA-AA) guidelines for the neuropathological assessment of AD^13^, which characterizes likelihood of AD according to an ’ABC’ staging. In this scheme amyloid plaques (’A’) are rated according to the method of Thal and colleagues^14^; neurofibrillary tangle severity is rated according to methods proposed by Braak and colleagues (’B’)^15, 16^; and neuritic plaques are rated according to the Consortium to Establish a Registry for Alzheimer’s Disease or “CERAD” (’C’) criteria^17^. ABC staging yields a rating for each case examined that corresponds to one of four categories: not AD, low AD, intermediate AD, or high AD neuropathological change (ADNC). In this study, similar to others that examined plasma biomarkers^18, 19^, our primary pathological grouping compared those classified as “high ADNC” compared with all other groups. Systematic Thal ratings for amyloid plaques were added later in the study and available for n=38 cases in the current sample, so our primary ADNC classification reflected a CERAD neuritic plaque rating of 2 (moderate) or 3 (severe), and a Braak rating of V or VI without incorporation of Thal ratings. In secondary analyses, we considered biomarker levels across the four degrees of AD neuropathological change among the n=38 participants with complete ADNC ratings (Thal, Braak, and CERAD).

### 2.5 Amyloid PET

A subset of participants from the clinical sample had previously undergone amyloid PET scanning with [^18^F]Florbetaben (8.1 mCi target dose). The standard uptake value ratio (SUVR) was calculated with data 50-70 minutes post-injection and an inferior cerebellar gray matter reference region in native space. All participants had high-resolution T1-weighted MRI and FreeSurfer^20^ was used to parcellate the brain and derive regions of interest (ROIs). A global composite SUVR was calculated as an average of frontal, temporal, and parietal cortex SUVRs, based on the FreeSurfer-defined ROIs. An SUVR cutscore of 1.25 was additionally used as an “amyloid positivity” threshold.

### 2.6 Analyses

We examined differences in each individual biomarker, includingAβ42/Aβ40, t-tau, NfL, P-tau181 and P-tau217, between clinically or pathologically-defined AD patients and controls and between PET amyloid positive and negative participants by t-tests. For autopsy data, we compared biomarker concentrations between those defined as high ADNC and all other groups with t-tests and examined receiver operating characteristics (ROC) with associated areas under the curve (AUCs) for diagnostic classification in the combined sample and then stratified by race/ethnicity. We used general linear models to examine differences in biomarker concentration across pathological ABC staging (not AD, low AD, intermediate AD, and high AD neuropathological change). In the clinical group, we used Pearson correlation and t-tests to test the association of each biomarker concentration with demographic features, including age, sex, APOE-ε4 status, and with amyloid PET SUVR as a continuous variable. We calculated AUCs for each biomarker with respect to diagnostic and amyloid status and examined differences across race/ethnicity groups. Prior to statistical analysis, frequency distributions were inspected for outlier values, which were removed from subsequent analyses. Three outliers were identified in the clinical group: one participant with a total tau concentration value of 472.3, one with a P-tau 181 concentration of 29.5, and one with NfL concentration of 628.5.

Finally, we used Cox regression analyses to examine the relationship between biomarker concentrations and AD diagnosis at the last available clinical follow-up visit among individual classified as controls at time of the first blood draw, using individuals who remained controls at last visit as reference. These analyses used the period between the initial blood draw and the last clinical diagnosis as the time-to-event adjusted for age at time of blood draw, sex, APOE-ε4, and race/ethnicity. We ran separate models for P-tau181 and P-tau217 as primary measures of phosphorylated tau.

## 3. Results

### 3.1 Participant characteristics

#### 3.1.1 Autopsy group

Average age at death and at last clinical follow-up were 88.63 (SD=6.80) years and 85.64 (SD=7.13) years, respectively (time interval between last evaluation/blood draw and autopsy, 2.99 [SD=2.77] years). Thirty-four (30%) had a clinical diagnosis of AD at their last WHICAP follow-up visit. Thirty-three (29%) had an ABC score indicating high neuropathological AD change. Table 1 displays demographic characteristics of the autopsy sample. Cases with high ADNC were similar in age and race/ethnicity distribution, but had a higher proportion of women and APOE-ε4 carriers than those with lower ADNC.

**Table 1.** Demographic and biomarker concentration characteristics in participants classified pathologically, clinically, and according to amyloid PET status. Note for the autopsy group, ’high AD’ refers to ADNC classification of high degrees of AD neuropathological change and controls include all other groups. For the clinical group, AD diagnosis is based on standardized clinical classification without consideration of biomarker status. For the PET subgroup, amyloid positivity was determined according to an SUVR threshold of 1.25.

|  |  | Pathological Status |  |  |  | Clinical Status |  |  |  | PET Amyloid Status |  |  |  |
| --- | --- | --- | --- | --- | --- | --- | --- | --- | --- | --- | --- | --- | --- |
|  |  | Controls | high AD | Total | Statistic | Controls | AD | Total | Statistic | - | + | Total | Statistic |
| N |  | 80 | 33 | 113 | - | 169 | 131 | 300 | - | 32 | 8 | 40 | - |
| Age, mean (SD) yrs |  | 84.93 (7.45) | 87.38 (6.04) | 85.64 (7.13) | t=1.67, p=0.09 | 81.01 (6.31) | 82.99 (6.49) | 81.87(6.46) | t=2.66 p=0.008 | 82.16 (5.19) | 84.25 (4.55) | 82.58 (5.08) | t=1.04, p=0.30 |
| Sex/gender, n(%) women | | 41 (51%) | 28 (85%) | 69 (61%) | $\chi^2=11.09$, p=0.001 | 109 (64%) | 91 (69%) | 200 (67%) | $\chi^2=0.82$, p=0.36 | 19 (59%) | 5 (63%) | 24 (60%) | $\chi^2=0.03$, p=0.87 |
| APOE, n (%) $\epsilon 4$ | | 16 (20%) | 13 (41%) | 32 (29%) | $\chi^2=4.89$, p=0.02 | 46 (27%) | 38 (29%) | 84 (28%) | $\chi^2=0.11$, p=0.73 | 11 (34%) | 6 (75%) | 17 (43%) | $\chi^2=4.32$, p=0.04 |
| Race/ethnicity, n (%) | White | 37 (46%) | 15 (45%) | 52 (46%) | $\chi^2=1.90$, p=0.38 | 63 (37%) | 37 (28%) | 100 (33%) | $\chi^2=2.71$, p=0.25 | 11 (34%) | 4 (50%) | 15 (38%) | $\chi^2=2.15$, p=0.34 |
|  | Black | 25 (31%) | 7 (21%) | 32 (28%) |  | 53 (31%) | 47 (36%) | 100 (33%) |  | 17 (53%) | 2 (25%) | 19 (48%) |  |
|  | Hispanic | 18 (25%) | 11 (33%) | 29 (26%) |  | 53 (31%) | 47 (36%) | 100 (33%) |  | 4 (13%) | 2 (25%) | 6 (15%) |  |
| Plasma biomarker concentration, mean (SD) pg/mL | Aβ42/Aβ40 | 0.06 (0.04) | 0.05 (0.01) | 0.06 (0.03) | t=1.24, p=0.21 | 0.05 (0.01) | 0.06 (0.01) | 0.05 (0.01) | t=0.57, p=0.56 | 0.06 (0.01) | 0.05 (0.005) | 0.06 (0.009) | t=0.83, p=0.40 |
|  | T-tau | 4.30 (3.22) | 4.12 (2.37) | 4.25 (2.98) | t=0.29, p=0.76 | 4.94 (2.13) | 4.90 (2.06) | 4.92 (2.09) | t=0.15, p=0.88 | 4.55 (2.13) | 5.64 (1.43) | 4.77 (2.04) | t=1.36, p=0.18 |
|  | p-tau 181 | 1.06 (0.81) | 1.93 (1.14) | 1.32 (1.00) | t=4.54, p<0.001 | 0.86 (0.73) | 1.24 (1.09) | 1.02 (0.92) | t=3.55, p<0.001 | 0.73 (0.61) | 1.35 (0.54) | 0.86 (0.64) | t=2.59, p=0.01 |
|  | p-tau 217 | 0.19 (0.16) | 0.51 (0.40) | 0.28 (0.29) | t=5.99, p<0.001 | 0.18 (0.17) | 0.32 (0.32) | 0.25 (0.26) | t=4.80, p<0.001 | 0.15 (0.14) | 0.39 (0.18) | 0.20 (0.18) | t=3.86, p<0.001 |
|  | NfL | 51.45 (51.11) | 34.41 (25.35) | 46.43 (45.63) | t=1.82, p=0.07 | 31.10 (28.96) | 36.55 (24.63) | 33.48 (27.25) | t=1.76, p=0.08 | 30.29 (20.73) | 30.31 (14.15) | 30.30 (19.43) | t<0.01, p=0.99 |

#### 3.1.2 Clinical group

Demographic characteristics at the time of the blood draw that was used to derive biomarker concentrations of participants included in the clinical cohort (n=300) are in Table 1. Compared with controls, those clinically diagnosed with AD were similar for sex distribution, race/ethnicity distribution, and APOE-ε4 allele frequency, but they were slightly older.

Of the individuals characterized as controls at time of blood draw, 71 subsequently developed AD. They were similar in age (t=0.52, p=0.46), sex (χ^2^<0.01, p=0.99), APOE-ε4 allele frequency (χ^2^=0.52, p=0.46), and race/ethnicity distribution (χ^2^=1.72, p=0.42) as those who remained controls. The last available diagnosis for these participants took place an average of 4.29 (SD=3.04) years after the blood draw used to derive biomarker concentrations.

#### 3.1.3 Amyloid PET subgroup

We compared demographic features between those classified as amyloid positive and negative (see Table 1). Amyloid positive individuals were more likely to be APOE-ε4 allele carriers, but were similar to amyloid negative participants in age, sex distribution, and race/ethnicity distribution. Fifty percent of amyloid positive individuals (n=4) were considered to have clinical AD at the diagnostic visit closest to the blood draw whereas 25% (n=8) amyloid negative individuals met clinical criteria for AD (χ^2^=1.90, p=0.16). PET scans were completed 3.79 (SD=20.8) months on average following the blood draw used to derive biomarker concentrations.

### 3.2 Biomarker concentration and neuropathological diagnosis

Individuals with high AD pathological changes had higher concentrations of P-tau 181 and P-tau 217, but similar Aβ42/Aβ40, T-tau, and NfL concentrations compared with those with lower amounts of AD pathological changes (see Table 1). P-tau 217 and P-tau 181 showed good diagnostic classification, but concentrations for the other biomarkers did not (see Table 2 and Figure 1). Although sample sizes were small and confidence intervals relatively wide, classification accuracy for P-tau 181 and particulary P-tau 217 concentrations was numerically better in non-Hispanic Black and Hispanic cases relative to Whites (see Table 2 and Figure 1). There was a monotonic increase in P-tau 181 (F=5.04, p=0.005) and P-tau 217 (F=6.27, p=0.002) but not the other biomarkers (F values range=0.12-0.25, p values range=0.70-0.94) across neuropathological classification groups (see Figure 2).

**Table 2.** Area under curve (AUC) statistics for each biomarker in the total sample and divided by race/ethnicity. 95% confidence intervals are presented in parentheses. Autopsy sample outcome is high AD pathology versus all other groups. Clinical outcome is clinical diagnosis of AD at time of blood draw used to derive biomarker concentrations.

|  |  | Total Group | non-Hispanic White | Black | Hispanic |
| --- | --- | --- | --- | --- | --- |
| Autopsy sample | Aβ42/Aβ40 | 0.41 (0.30-0.52) | 0.36 (0.20-0.53) | 0.51 (0.29-0.72) | 0.38 (0.17-0.58) |
|  | t-tau (pg/mL) | 0.51 (0.40-0.63) | 0.44 (0.27-0.61) | 0.41 (0.20-0.62) | 0.67 (0.45-0.88) |
|  | P-tau181 (pg/mL) | 0.77 (0.67-0.87) | 0.65 (0.48-0.82) | 0.94 (0.85-1.00) | 0.81 (0.66-0.97) |
|  | P-tau217 (pg/mL) | 0.83 (0.75-0.92) | 0.75 (0.61-0.89) | 0.96 (0.90-1.00) | 0.84 (0.68-1.00) |
|  | NfL (pg/mL) | 0.40 (0.30-0.52) | 0.36 (0.21-0.50) | 0.45 (0.19-0.72) | 0.43 (0.21-0.66) |
| Clinical sample | Aβ42/Aβ40 | 0.50 (0.43-0.56) | 0.50 (0.38-0.63) | 0.51 (0.40-0.63) | 0.46 (0.35-0.58) |
|  | t-tau (pg/mL) | 0.50 (0.44-0.57) | 0.53 (0.42-0.65) | 0.55 (0.44-0.67) | 0.41 (0.30-0.53) |
|  | P-tau181 (pg/mL) | 0.60 (0.54-0.67) | 0.69 (0.59-0.80) | 0.62 (0.51-0.74) | 0.51 (0.40-0.64) |
|  | P-tau217 (pg/mL) | 0.63 (0.57-0.70) | 0.71 (0.60-0.82) | 0.67 (0.57-0.78) | 0.52 (0.40-0.64) |
|  | NfL (pg/mL) | 0.58 (0.52-0.66) | 0.65 (0.53-0.76) | 0.64 (0.53-0.76) | 0.51 (0.40-0.63) |

**Figure 1.**
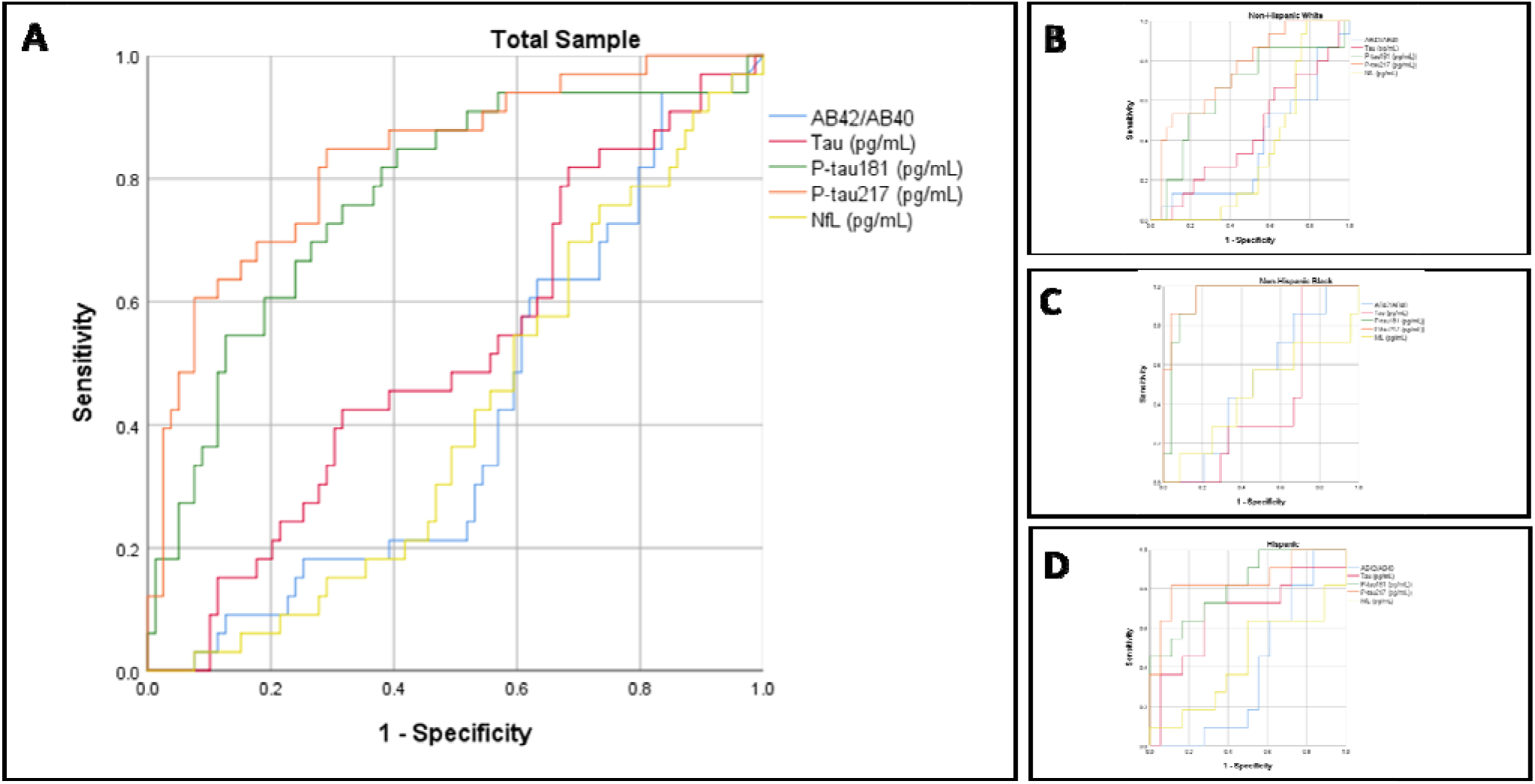
Receiver operating curves for classification of postmortem diagnosis of AD. Panel A shows the total sample. Panel B shows non-Hispanic Whites. Panel C shows non-Hispanic Blacks. Panel D shows Hispanics.

**Figure 2.**
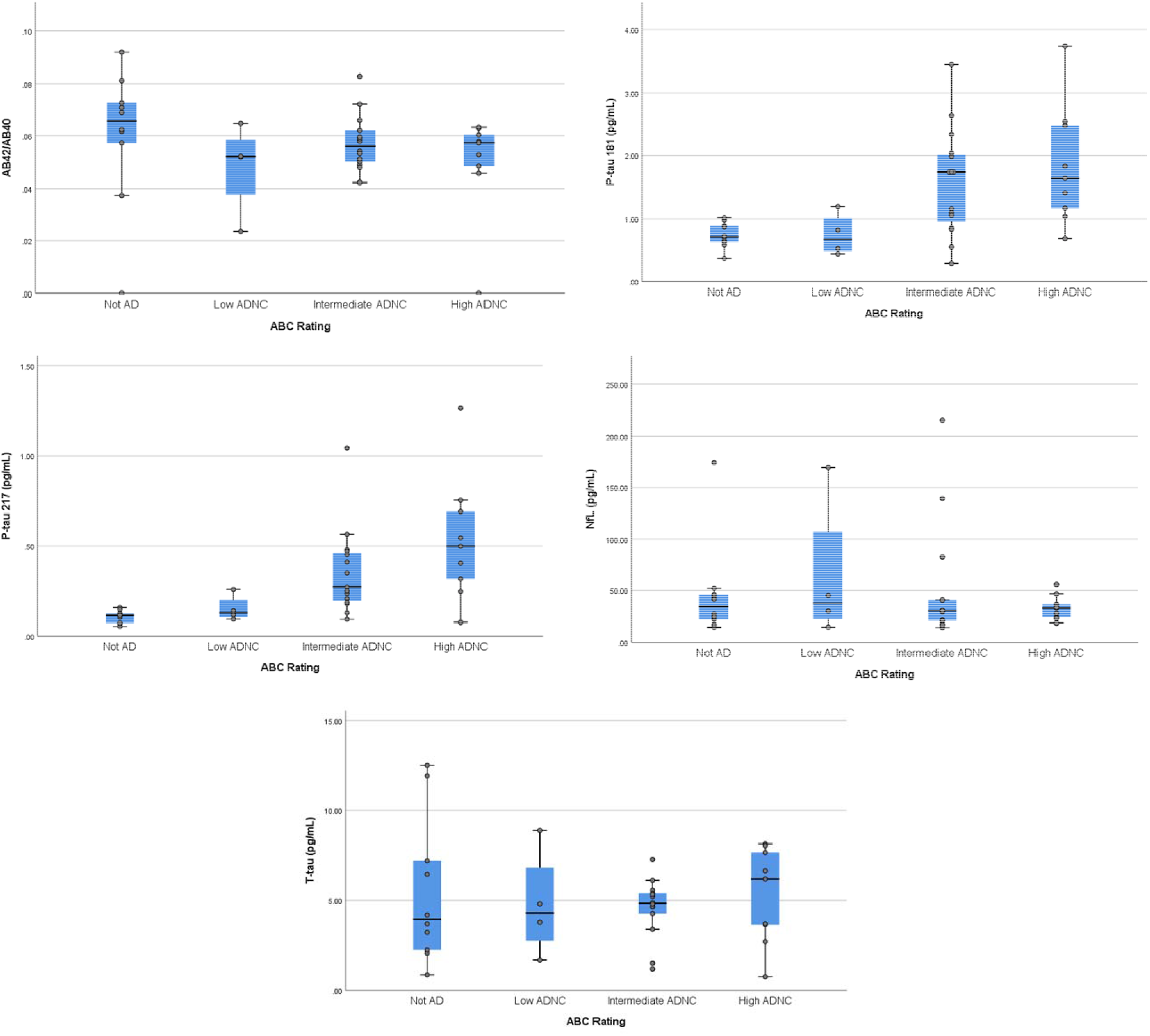
Biomarker concentrations across pathological ’ABC’ ratings. Midline represents median, box represents 25^th^ and 75^th^ percentile, and T-bars represent 95% confidence interval. Individual subject data points are superimposed.

**Figure 3.**
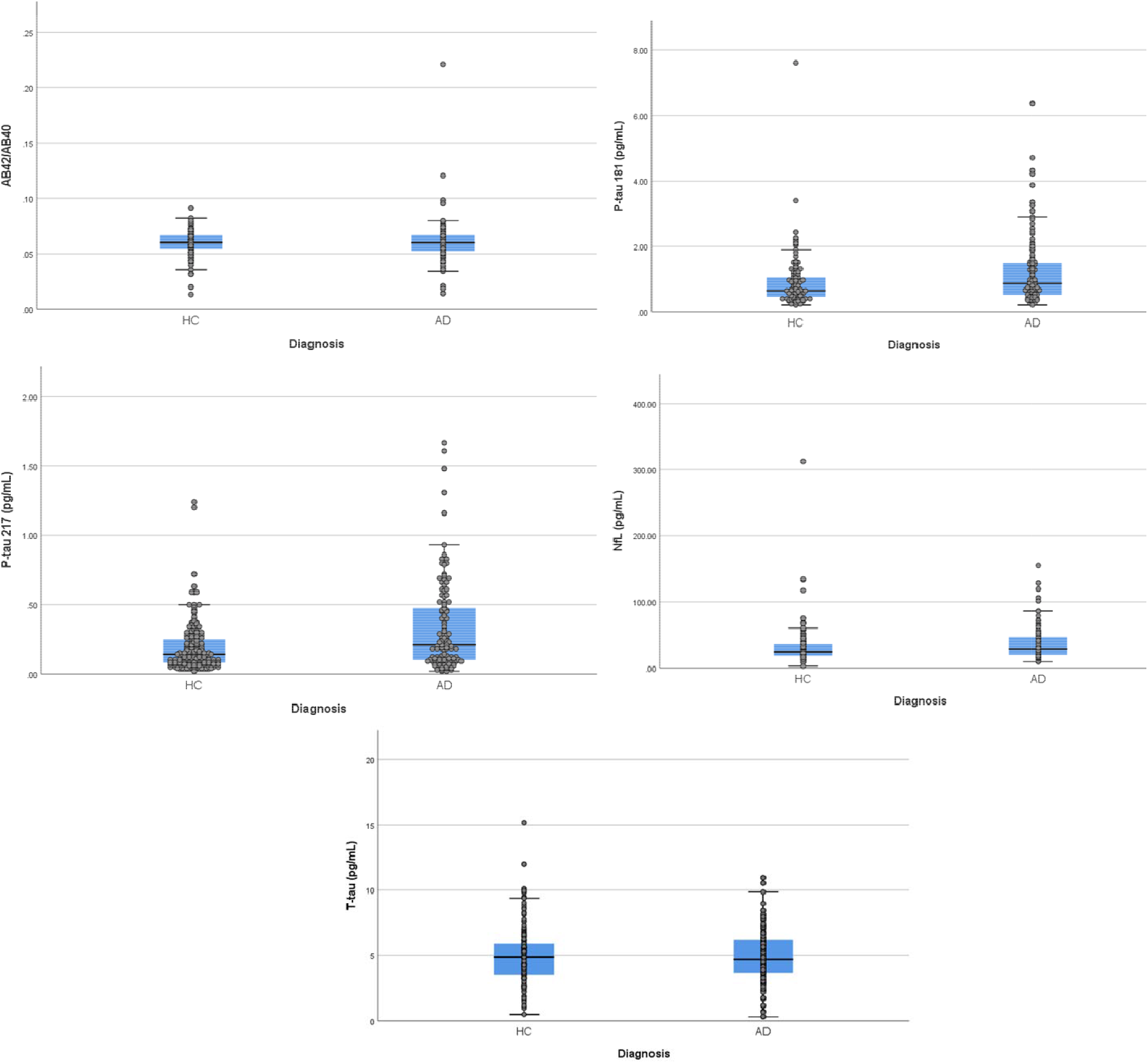
Differences between clinically diagnosed patients with AD and HCs in absolute concentrations of each plasma biomarker. Midline represents median, box represents 25^th^ and 75^th^ percentile, and T-bars represent 95% confidence interval. Individual subject data points are superimposed.

### 3.3 Biomarker concentration and clinical characteristics

#### 3.3.1 Demographic features

Increased age was associated with Aβ42/Aβ40 (r=-0.099, p=0.08), t-tau (r=0.149, p=0.01, P-tau 181 (r=0.214, p<0.001), P-tau 217 (r=0.192, p=0.001), and NfL (r=0.286, p<0.001) concentrations. Biomarker concentrations did not differ between men and women (t value range= 0.14-1.12, p value range=0.25-0.88). Compared with non-carriers, APOE-ε4 allele carriers had higher concentrations of P-tau181 (mean±SD=1.20±0.95 vs. 0.96±0.91, t=2.03, p=0.04) and P-tau217 (mean±SD=0.31±0.23 vs. 0.22±0.26, t=2.78, p=0.006), but were similar for all other biomarker concentration levels (t value range 0.16-1.30, p value range 0.86-0.19). Biomarker concentrations were similar across the three race/ethnicity groups (F value range 0.61-2.15, p value range 0.54-0.11).

#### 3.3.2 Clinical diagnosis at time of blood draw

We examined the differences in biomarker concentrations between participants with clinically diagnosed AD and controls(Table 1 and Figure 4). Participants with AD had higher concentrations of P-tau 181 and P-tau 217and higher NfL levels, and similar Aβ42/Aβ40 and t-tau levels compared with controls. We constructed ROCs for each biomarker as a function of diagnostic status (Figure 4), which revealed the best diagnostic classification for P-tau concentrations (Table 2). When stratified by race/ethnicity, classifications were comparable among non-Hispanic White and Black participants but AUCs among Hispanic participants were relatively low for all biomarkers measured (Table 2 and Figure 4).

**Figure 4.**
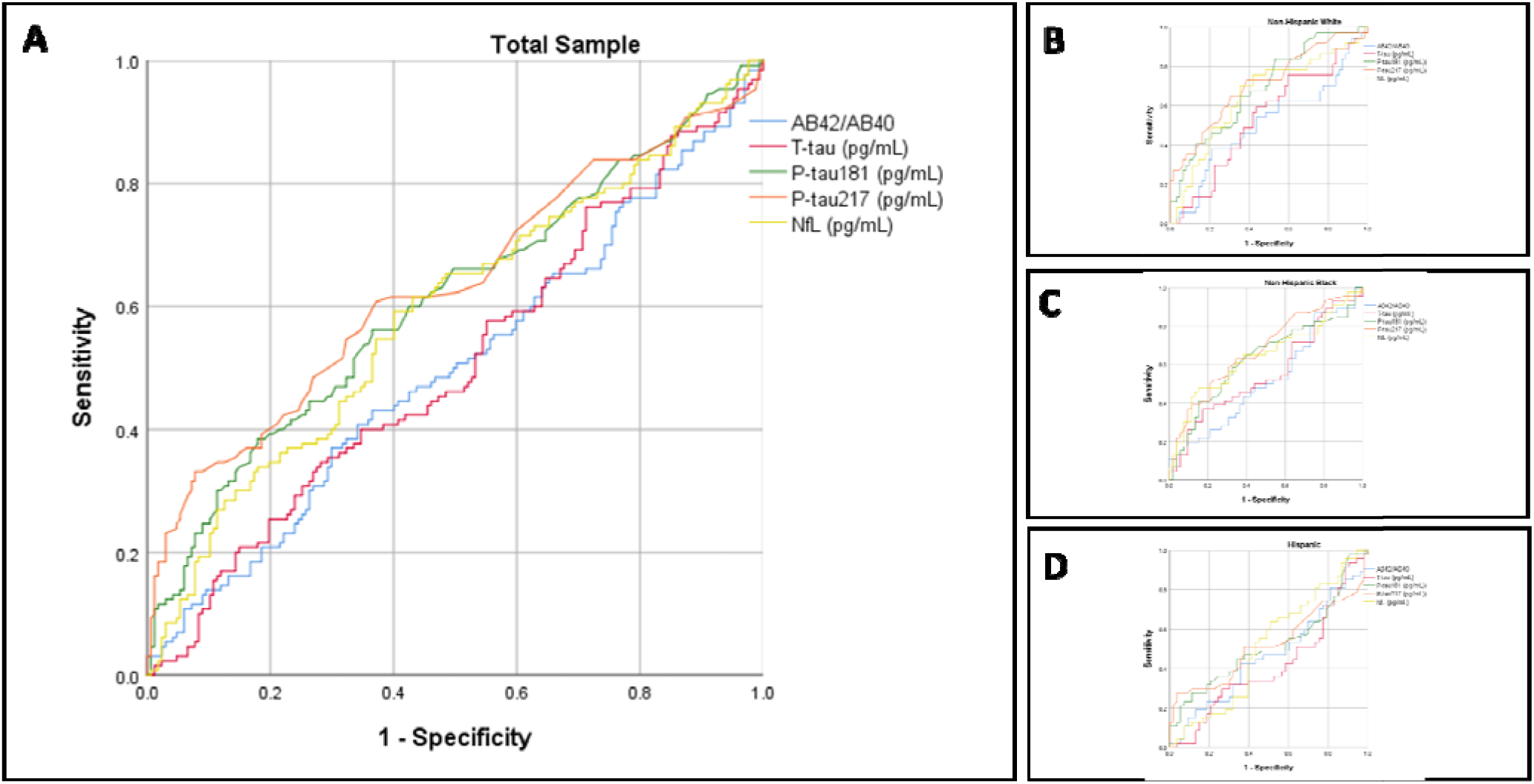
Receiver operating curves for classification of clinical diagnosis of AD. Panel A shows the total sample. Panel B shows non-Hispanic Whites. Panel C shows non-Hispanic Blacks. Panel D shows Hispanics.

#### 3.3.3 Amyloid PET

Compared with amyloid negative individuals the amyloid positive individuals had higher levels of P-tau 217 and P-tau 181 (Table 2). Total tau concentrations trended higher in amyloid positive individuals, but Aβ42/Aβ40, and NfL levels were quite similar between the groups. These group differences reflect what was observed when considering amyloid PET cortical SUVR as a continuous measure. There were strong correlations of amyloid SUVR with P-tau 217 (r=0.48, p=0.002) and P-tau 181 (r=0.36, p=0.02), more modest associations with T-tau (r=0.23, p=0.15) and Aβ42/Aβ40 (r=-0.16, p=0.30), and trivial correlations with the other biomarkers (r value range = 0.01-0.07, p value range = 0.93 – 0.65). When we examined classification of amyloid positivity with ROCs, the findings paralleled the other results: AUCs were greatest for P-tau 217 (AUC:0.84, 95% CI:0.68-0.99) and P-tau 181 (AUC: 0.82, 95%CI:0.65-0.99); AUCs for the other biomarkers ranged from 0.40 (95%CI: 0.22-0.59; Aβ42/Aβ40) to 0.56 (95%CI: 0.34-0.79; NfL). We did not examine differences across race/ethnicity groups because of the small sample and low number of amyloid positive participants.

### 3.4 Risk of subsequent clinical AD among those without dementia at the first blood draw

The mean and median for Aβ42/Aβ40 ratio were identical, but the P-tau 181 and P-tau217 levels were skewed. Therefore, we used the median value of these biomarkers to estimate the risk of developing AD. The model indicated an overall effect of biomarker concentration on subsequent development of clinical AD was similar for P-tau 181 (χ^2^=22.9, p<0.002) and P-tau217 (χ^2^=20.8, p<0.007) in independent analyses. We found reduced Aβ42/Aβ40 ratio and an increase in either P-tau217 or P-tau181 to be associated with increased risk of developing AD (Table 3). None of the other biomarker concentrations were associated with increased risk, but APOE-ε4 allele was associated with a slightly higher risk.

**Tables 3a and 3b.** The association between plasma biomarker concentrations derived at blood draw and the subsequent clinical AD diagnosis approximately 4 years later. We used Cox Regression in which the time to event was calculated as the period between the blood draw and last diagnostic assessment. Biomarker predictors were dichotomized according to their median; outcome was coded as 1=healthy control 2=incident AD and we adjusted for age at last visit, sex (reference group is male sex), ethnic group and APOE-e4, Table 3a shows the adjusted results using P-tau 181, while table 3b displays the adjusted results using P-tau 217.

| Table 3a |  | B | p | OR (95% CI) |
| --- | --- | --- | --- | --- |
| P-tau 181 (pg/mL) |  | 0.85 | 0.002 | 2.3 (1.4-4.0) |
| Aβ42/Aβ40 | | 0.627 | 0.01 | 1.9 (1.1-3.2) |
| T-tau (pg/mL) |  | -0.06 | 0.28 | 0.94 (0.85-1.4) |
| NfL (pg/mL) |  | 0.005 | 0.19 | 1.01 (0.99-1.0) |
| Age |  | 0.064 | 0.02 | 1.07 (1.01-1.12) |
| APOE e4 allele |  | -0.233 | 0.43 | 0.79 (0.44-1.4) |
| Race/<br>ethnicity | Non-Hispanic<br>White | 1.0 | 0 | reference |
|  | Black | -0.106 | 0.75 | 0.9 (0.47-1.87) |
|  | Hispanic | 0.13 | 0.69 | 1.14 (0.61-2.1) |
| Sex |  | -0.412 | 0.13 | 0.66 (0.38-1.14) |

| Table 3b |  | B | p | OR (95% CI) |
| --- | --- | --- | --- | --- |
| P-tau217 (pg/mL) |  | 0.67 | 0.04 | 1.96 (1.1-3.3) |
| Aβ42/Aβ40 | | 0.642 | 0.016 | 1.9 (1.1-3.2) |
| T-tau (pg/mL) |  | -0.03 | 0.57 | 0.97 (0.88-1.1) |
| NfL (pg/mL) |  | 0.005 | 0.196 | 1.01 (0.97-1.01) |
| Age |  | 0.058 | 0.036 | 1.06 (1.0-1.1) |
| APOE e4 allele |  | -0.254 | 0.39 | 0.78 (0.43-1.4) |
| Race/<br>ethnicity | Non-Hispanic<br>White | 1.0 | - | Reference |
|  | Black | -0.064 | 0.85 | 0.94 (0.49-1.8) |
|  | Hispanic | 0.046 | 0.089 | 1.04 (0.55-2.0) |
| Sex |  | -0.36 | 0.19 | 0.69 (0.41-1.2) |

## 4. Discussion

We found that the plasma biomarker concentrations of phosphorylated tau, particularly P-tau 217, were strongly associated with measures of AD pathology, and were the best predictors of autopsy-confirmed AD. As expected, these observations did not completely translate to clinically-diagnosed participants due to known limitations of clinical diagnosis for AD pathology where nearly half of early symptomatic patients can have no AD pathology if evaluated by PET. Nonetheless, at the time of blood draw those diagnosed clinically with AD had similar plasma biomarker profiles to the participants who were ultimately autopsy confirmed AD, including higher P-tau 217 and NfL concentrations. P-tau concentration differences most reliably classified clinical diagnostic groups at the time of the blood draw. Similarly, P-tau biomarkers were also associated with PET markers of amyloid pathology, more so than other plasma biomarkers, including Aβ42/Aβ40. Among individuals classified as controls at time of blood draw, lower Aβ42/Aβ40 ratio and higher P-tau 217 or P-tau181 concentrations were associated with increased risk of subsequent AD diagnosis approximately four years later. Blood-biomarkers^19^ can change as early as 10 years prior to the onset of dementia, making follow up time an important consideration. In our study, the average time between blood draw and the most recent diagnostic visit was about four years, but within this time frame both the amyloid and P-tau measures contributed to the risk of subsequent AD among participants without dementia at baseline.

Plasma levels of P-tau 217 consistently outperformed all other biomarkers across analyses, achieving with AUCs of 0.83 overall, and 0.84 and 0.96 in Hispanic and Black participants with pathology data. However, in the analysis of those who converted from controls to clinical AD, both P-tau 181 and P-tau 217 as well as the Aβ42/Aβ40 ratio were the most robust predictors of change. These observations are very consistent with a recent study that showed similar classification in three separate cohorts^19^. It is clear that among the currently-available plasma biomarkers, P-tau 217 concentrations reflect underlying tau pathology with the greatest fidelity and are useful to aid the clinical diagnosis of AD. As expected, and due to the association of age and APOE ε4 allele status with AD, biomarker concentrations were somewhat age-dependent and varied with respect to APOE ε4 allele status. In general, the concentrations did not differ across race/ethnicity or sex, although larger studies will be necessary to understand fully the potentially moderating effects of these demographic variables.

The results we observed for P-tau181 are similar to what was observed by Janelidze et al^7^, but only when we dichotomized the results around the median and we observed similar findings to P-tau217. P-tau217 and P-tau181 did not differ as predictors of progression in this study, even when dichotomized. These differences when compared with Janelidze et al^7^, may be attributed to the older average ages in this study and higher likelihood of the presence of comorbidities, which are age dependent^21^. Additionally, Janelidze et al.^7^ separated AD from other forms of dementia, aiding in the specificity of P-tau. For this study, we used strict criteria to select individuals with clinical AD and did not include other forms of dementia.

The WHICAP cohort is racially/ethnically diverse and community-based. Most previous studies that incorporated plasma biomarkers included clinic-based samples and participants with minimal racial and ethnic diversity. Here, we provide preliminary evidence that plasma biomarker data can be incorporated successfully into community-based research, correspond with neuropathological changes seen in AD, and perform equally well or better in racial/ethnic groups typically under-represented in aging research.

While the observations shown in this study highlight the promise and potential of plasma-based biomarkers in identification of AD pathology and risk of developing symptoms of dementia, they fall short as stand-alone diagnostics due to the variability between signs of pathology and symptoms of AD.. Based on the findings here, P-tau 181 or P-tau217 would augment the clinical diagnostic accuracy of AD compared with other forms of dementias, as well as the presence of pathology preceeding detectable symptoms or existing as a secondary co-pathology. This issue may be addressed with new tools to detect cognitive symptoms earlier and with greater specificity to a particular pathology.

Although the plasma biomarkers tested showed associations with clinical dementia, only the P-tau associations improved when confirmed either with PET or postmortem examination. There are at least two potential explanations for these observations. First, pathological and PET diagnoses are more definitive and there is always some diagnostic imprecision with clinical diagnoses alone. Second, and related, clinical AD is a heterogeneous condition, determined by multiple pathologies whose unique combination may vary across individuals^22^. While much improved over previous generations of plasma biomarkers, current biomarkers require longitudinal studies to understand their temporal dynamics with respect to timing of neuropathological changes as well as clinical disease progression.

Prior research on AD biomarkers across racial/ethnic groups has been challenged by small sample sizes and selection biases and has mostly focused on comparing non-Hispanic Whites to Blacks, while inclusion of Hispanics is less frequent^23-30^. Large population studies are required to provide key information as to the role race/ethnicity has in disease prevalence or the socioeconomic factors that contribute to dementia. This study provides evidence that P-tau217 may be useful as an indicator of pathology that will aid in the evaluation of other biomarkers as well as understanding race/ethnicity and socioeconomic factors in the context of large community-based studies.

Taken together the results here provide encouraging data for the use of blood-based biomarkers in diverse cohorts across clinical settings and in observational, epidemiological studies. Establishing a universal cut-point for P-tau 181 or P-tau 217 should be a priority in future studies of these biomarkers.

## Data Availability

WHICAP data are available through written request to the corresponding author.

## Acknowledgments

Data collection and sharing for this project was supported by the Washington Heights-Inwood Columbia Aging Project (WHICAP; P01AG07232, RF1AG054023, R01AG037212, R56AG034189, R01AG034189, R01AG054520) funded by the National Institute on Aging (NIA). This manuscript has been reviewed by WHICAP investigators for scientific content and consistency of data interpretation with previous WHICAP Study publications. We acknowledge the WHICAP study participants and the WHICAP research and support staff for their contributions to this study.

Florbetaben was provided to the study by Life Molecular Imaging (formally Piramal Imaging) under an Investigator Initiated Study.

